# The impact of Seasonal Malaria Chemoprevention and relationship to parasite drug resistance in a nomadic pastoralist community in Northern Kenya

**DOI:** 10.64898/2025.11.30.25341333

**Authors:** Beatrice Rotich, Emmah Kimachas, Joseph Kipkoech, Erastus Kirwa, Christine Markwalter, David Ekai, Ann Mwangi, Suzanne Van Hulle, Diana Menya, Wendy Prudhomme O’Meara

## Abstract

**Introduction:** Following WHO’s updated guidelines for SMC, countries beyond the Sahel region are exploring the suitability of SMC to protect children in areas with sharply seasonal transmission. Sulphadoxine-pyrimethamine with amodiaquine (SPAQ) is the standard regimen, leading to concerns about the efficacy of SMC in areas with higher levels of SP resistance. Here we report the impact of an SMC program implemented in Turkana Central, an area with moderate-to-high SP resistance. We examine the relationship between parasite resistance and breakthrough infections in children receiving SMC.

**Methods:** We conducted a parallel cohort study during the 2024 peak malaria season (June - October) with 10 randomly-selected villages in the SMC delivery subcounty (Turkana Central) and 10 from the adjacent subcounty (Turkana North). Community Health Promoters conducted weekly surveillance of malaria symptoms. Symptomatic children were tested with a Rapid Diagnostic Test and blood spot samples were collected. Using Poisson regression, an incidence rate ratio was used to measure SMC efficacy where the primary outcome was malaria-like symptoms with a positive diagnostic test. *Pf* positive samples from symptomatic infections were tested for SP resistance alleles within the *Pfdhfr* and *Pfdhps* genes.

**Results:** 398 children were followed from 2 weeks prior to SMC until four weeks after the final cycle. In Turkana Central, 145/198 (73%) children received complete SMC. There was a 71% reduction in malaria incidence attributable to SMC (Adjusted IRR 0.29, 95% CI:0.13-0.67). There were 156 *Pf-*positive samples collected during active surveillance of which 68 were fully characterized at all three codons. The proportion of infections with mutations in the *Pfdhps* gene was significantly higher in children receiving SMC and increased with increasing cycles of SMC received.

**Conclusion:** SMC with SPAQ is highly effective for preventing malaria in areas with moderate prevalence of SP resistance. Despite this, infections in children who received SMC were more likely to harbor resistant parasites.

## Background

Seasonal Malaria Chemoprevention (SMC) has been used in the Sahel for more than a decade to prevent morbidity and mortality in vulnerable populations. It involves the administration of antimalarials that are not part of local first or second line treatment of malaria to children in order to protect them from malaria (1,2). The most commonly used SMC regimen is Sulphadoxine-Pyrimethamine (SP, an antifolate drug) and Amodiaquine (AQ) which are administered in therapeutic doses at 28 day intervals, throughout the high transmission period, usually 3-5 months. Amodiaquine clears existing infections while SP, due to its long half-life, ensures a prolonged antimalarial effect on new infections. There is substantial evidence from randomized studies and large scale implementation showing SMC with SP-AQ to be highly effective, safe, and cost-effective for the prevention of malaria among children in areas with seasonal malaria transmission (3–6).

Following WHO’s updated guidelines to consider scale up of SMC beyond the Sahelien region, countries in East Africa (Uganda, Sudan and Kenya) are now exploring SMC as part of their prevention arsenal, targeting areas with highly seasonal transmission. However, in East Africa, parasites harbor higher levels of SP resistance than those found in West Africa (7,8). In Kenya, the use of SP for treatment of clinical malaria was stopped in the mid-2000s as a consequence of high levels of treatment failure (9) and studies have documented the persistence of antifolate resistance markers despite stopping clinical use (7,10). Still, there is evidence to support the continued use of SP for prevention of malaria in areas with moderate levels of antifolate resistance, for example in pregnancy (Intermittent Preventive Treatment for malaria in pregnancy, IPTp) (11–13). A recent SMC pilot in Uganda, where antifolate resistance is high, demonstrated a 90% reduction in malaria cases (14). However, the relationship between antifolate (SP) resistance and efficacy of chemoprevention with SP-AQ is not well understood.

Here we report the effectiveness of the first SMC program in Kenya implemented in a region where resistance to antifolates has persisted despite the absence of IPTp and the removal of SP from clinical case management. Communities in this region are highly mobile, sleep out in the open, and have poor access to formal health services. These factors result in low penetration of conventional malaria case management or vector control tools such as insecticide-treated bed nets and indoor residual spraying. Using active case detection to measure incidence of malaria in parallel cohorts of children in the intervention and non-intervention communities, we document the effectiveness of SMC using SPAQ under routine implementation. We sampled parasites during incident infections in both groups and tested for the presence of antifolate resistance mutations in order to directly measure the relationship between SMC effectiveness, breakthrough infections, and parasite resistance.

## Methods

### Study area and SMC program

Turkana County is located in the far northwest corner of Kenya and is bordered by Uganda to the west and South Sudan and Ethiopia to the north. It is the largest county in Kenya, and covers 13% of the country by area. The county is home to 1.2 million people, 13% of whom are under 5 years of age.

Most areas have limited access to basic infrastructure such as roads, health facilities and clean water (15).

The climate is characterized by extreme weather conditions. It receives rainfall between 120mm and 500mm annually, erratic in distribution and timing, with mean annual temperature ranging between 26°C and 38°C. Many families in Turkana are semi-nomadic, moving with their herds in search of pasture due to the small amounts of rainfall received each year. Mean temperatures in Turkana are rising sharply from year to year (16). Malaria outbreaks occur frequently (17,18) with the highest upsurge in a 2024 outbreak during which 400,000 cases were recorded in health facilities in the county (D. Ekai, personal communications).

Turkana was previously considered to be largely outside of the malaria risk map, but recent studies have revealed endemic transmission in the region, despite receiving little rainfall (19–21). According to health facility records, approximately 80% of cases occur within a span of 5 months and 30% percent of all malaria cases occur in children under 5 years.

SMC was implemented in Turkana Central sub county by Turkana County Ministry of Health with support from Catholic Relief Services (CRS) during the 2024 high transmission season in 5 cycles at 28 day intervals from June 2024 to October 2024. SMC was primarily delivered door to door by Community Health Promoters (CHPs) supplemented by mobile fixed points and outreach programs adapted to the remote and mobile population. The programme targeted children 3-59 months old. More than 40,000 children were reached with at least one cycle of SMC.

### Study design and sample size

A prospective, partially-randomized parallel cohort study was conducted to determine the impact of SMC on malaria incidence. The intervention was not randomized, but the cohort villages were randomly selected in order to be as representative as possible. A cohort of children age 6-59 months was established in the intervention area (Turkana Central) and the comparison area in the neighboring Turkana North. The intervention cohort was enrolled from Turkana Central by randomly selecting two villages from each of the five wards from a full list of 644 villages. Within each ward, villages were selected with probability proportional to size. Random selection was constrained in Turkana North non-intervention area to 47 villages corresponding to four health facilities that had similar malaria test positivity rates to facilities linked with selected Turkana Central villages.

Household listings for selected villages were obtained and augmented through cross-checking community health registers, either a paper Ministry of Health register (MOH 513) or the electronic health register (eCHIS). A random sample of 20 households was selected through systematic random sampling at each village. A child was eligible if they were between 6-59 months and would not pass their fifth birthday by the start of SMC in mid-June. If more than one child was eligible in a household, an auto-populated randomized allocation unrelated to household ordering was used for selection of children. In villages with fewer than 20 eligible children, we enrolled an additional child from a subset of households.

A sample size of 200 children across 10 clusters in the intervention area and non-intervention area was sufficient for this study to have a desired 85% statistical power at 5% level of significance to measure a 50% difference in the proportion of children who have at least one malaria episode. We conservatively estimated that 40% of children in the intervention area would have malaria during the 5 month follow up period.

### Outcome measures

The primary outcome was a positive malaria diagnostic test from any source accompanied by malaria-like symptoms (specifically, fever, history of fever, aches, chills, lethargy). The comparison of interest was the incidence of malaria in the group receiving SMC compared to the group not receiving SMC. The secondary outcome was incidence of any malaria-like illness regardless of whether they had a test or not.

Illnesses were documented through both active and passive surveillance. CHPs visited households approximately once per week to ask about fever and illness. They offered RDT testing at the home when an enrolled child was unwell. Children with a positive test were referred for treatment at the local health facility. The household was encouraged to contact the CHP whenever the child had symptoms of malaria to receive a free RDT. A DBS was prepared each time a CHP conducted an RDT during a household visit. The number of cycles of SMC received were also documented. In the event that the child was unwell and the CHP could not visit or the child was taken to a health care provider without contacting the CHP, the instances of illness or treatment seeking was documented at the next weekly visit. The child’s health card or medical booklet was reviewed to confirm details of the health facility visit when possible. Only illness accompanied by a positive diagnostic test from the CHP or the facility were included for the primary outcome.

### Data collection and analysis

At enrollment, field researchers captured household and child data directly into a tablet-based form using Redcap. CHPs conducted routine follow-up visits and completed paper questionnaires which were then entered into the Redcap database. At the end of the SMC period (28 days after the final cycle of SMC) all households participating in the cohort were visited by the study team. On this final visit, a brief tool developed in KoboCollect Android App (https://www.kobotoolbox.org/) captured data on the child’s travel over the five month follow up period, child illness during the final month of follow up and information on child’s participation in the SMC programme (in intervention area only). Details were confirmed by reviewing government-issued SMC cards when available.

This was an intention-to-treat analysis. A mixed effects Poisson regression model was used to estimate the incidence rate ratio of malaria episodes in the treatment versus comparison group. Models were adjusted for pre-specified covariates: gender and age of the child, whether the child had a bed net for their sleeping space, whether the child slept outside and the village-level prevalence of malaria infection at baseline. Data were analyzed in Stata v.18 (StataCorp, College Station TX) and Rstudio v2025.05.0+496 (22)with R v4.3.1 (23) using the following packages: tidyverse v2.0.0(24), lme4 v1.1-34(25), survival v3.8-3 (26), survminer v0.5.0 (27), broom v1.0.7 (28), and kableExtra v1.4.0 (29)

### Samples and laboratory analyses

At enrollment and each instance of an RDT performed by the CHP, dried blood spot (DBS) samples were collected from participants. DBS were prepared with 5-10ul of capillary blood from a finger prick, which was dropped onto a filter paper and allowed to dry completely. Filter papers were stored in plastic zip lock bags with desiccant until transportation to the laboratory.

Baseline and active case detection samples were tested for the presence of *P. falciparum* genomic DNA by a nested PCR beginning with an amplification using Phusion Blood Direct PCR Kit (Thermoscientific) with Plu1/5 primers described by Snounou and Sing (30). Briefly, a 3mm punch of the filter paper is suspended in 40ul of mastermix (1x Phusion Blood PCR Buffer, 0.5uM forward and reverse primer, 1.6 Units of Phusion Blood II DNA Polymerase), denatured at 98°C for 5 minutes then and cycled 35 times at 98°C for 1 second, 60°C for 5 seconds and 72°C for 30 seconds. A second round of amplification was based on previously described primers (31) as follows: 1ul of the primary reaction as template and 0.25uM forward and reverse primers, 0.3uM probe, and 1x SSO Advanced Universal Probes Supermix (Bio-rad).

*P*.*falciparum* positive DBS were further processed as follows: genomic DNA was extracted from a fresh 5mm punch of the DBS using a chelex-based protocol(32) and samples were subjected to further testing for genetic markers of resistance to sulphadoxine pyrimethamine. An allelic discrimination assay was employed based on Alker *et al* (33). There are five dominant mutations in the *dhfr* and *dhps* genes that have been shown to correspond to parasite antifolate resistance in East Africa: *dhps* A437G and K540E, *dhfr* N51I, C59R and S108N. Substitutions at dhfr N51I and S108N are at fixation in Kenya and have remained so more than 15 years after the withdrawal of SP (34). Therefore, we focused our analysis on the remaining three substitutions which are most likely to show heterogeneity in the local parasite population: *dhps* A437G, *dhfr* C59R, and *dhps* K540E. Each reaction contained 5 microliters of chelex extracted genomic DNA, 0.3uM forward and reverse primers, 1x SSO Advanced Mastermix and 0.2uM of each of two probes, one corresponding to the mutant and the other to the wildtype allele. Fluorescence signals from either probe or both identified infections harboring wild-type, mutant or mixed infections at that position.

### Ethics and Informed Consent

Written informed consent was obtained from the parents or guardians of children participating in this study. Approval for this study was granted by Moi University Institutional Research and Ethics Committee (IREC) and Duke University Institutional Review Board. In addition, PATH research committees reviewed the study protocol on behalf of the project funder, President’s Malaria Initiative (PMI). This study was registered with Pan African Clinical Trials Registry (PACTR), Trial Number PACTR20240570172.

### Participant and public involvement

Participants were not directly involved in the design of this study. However, the input and information provided by the public and community leadership during the formative research phase(35) was critical for designing the SMC programme. Ministry of Health and community leadership were involved in all aspects of the study design, randomization, enrollment and supervision of study activities.

## Results

### Study population

398 eligible children were enrolled in 389 households, 198 children in Turkana Central (intervention area) and 200 in Turkana North (comparison area), respectively. Nine households had more than one child participating in the cohort. The two cohorts were similar at baseline (Table 1) – 49% were female, the mean age was 28.9 months. ITN ownership was low, only 50% of children had a net for their sleeping space. 68 (17%) children reported symptoms on the day of enrollment and were tested with an RDT, of whom 19% were positive (n=13, 4 in Turkana North and 9 in Turkana Central). Complete household information is available in Table S1.

**Table 1:** Characteristics of participants at enrollment.

|  | **Overall**<br>N = 398<sup>1</sup> | **Turkana Central**<br>N = 198<sup>1</sup> | **Turkana North**<br>N = 200<sup>1</sup> |
| --- | --- | --- | --- |
| **Age (months)** | 28.9 (14.3) | 28.7 (14.5) | 29.1 (14.2) |
| **Proportion female** | 195 (49%) | 96 (48%) | 99 (50%) |
| **Has bednet for sleeping space** | 198 (50%) | 106 (54%) | 92 (46%) |
| **Sleeps outside** | 88 (22%) | 40 (20%) | 48 (24%) |
| **Symptomatic at enrollment** | 68 (17%) | 38 (19%) | 30 (15%) |
| **RDT positive with symptoms** | 13 (3.3%) | 9 (4.5%) | 4 (2.0%) |

<sup>1</sup>Mean (SD); n (%)

**Table 2:** Illness events, follow-up time and SMC coverage in the cohort participants.

| **Characteristic** | **Overall**<br>N = 398<sup>1</sup> | **Turkana Central**<br>N = 198<sup>1</sup> | **Turkana North**<br>N = 200<sup>1</sup> |
| --- | --- | --- | --- |
| **Febrile cases** | 566 | 383 | 183 |
| **Malaria cases** | 207 | 149 | 58 |
| **Had >1 malaria episode** | 47 | 36 | 11 |
| **Remained malaria-free** | 258 | 99 | 159 |
| **Lost to follow-up** | 23 | 9 | 14 |
| **SMC coverage** |  |  |  |
| Complete | - | 143 (72.2%) | - |
| Partial | - | 39 (19.7%) | - |
| None | - | 2 (1.0%) | - |
| Unknown |  | 14 (7.1%) |  |
| **Person months of observation** | 2,179 | 1,128 | 1,051 |

<sup>1</sup>Mean (SD); n (%)

Parasite prevalence was measured by PCR in all children at enrollment and varied between villages (Figure 2). The prevalence of parasite infection overall was 0.20 (95%CI: 0.16-0.24), with slightly lower prevalence in Turkana North (0.16 vs. 0.23, p=0.077)

**Figure 1:**
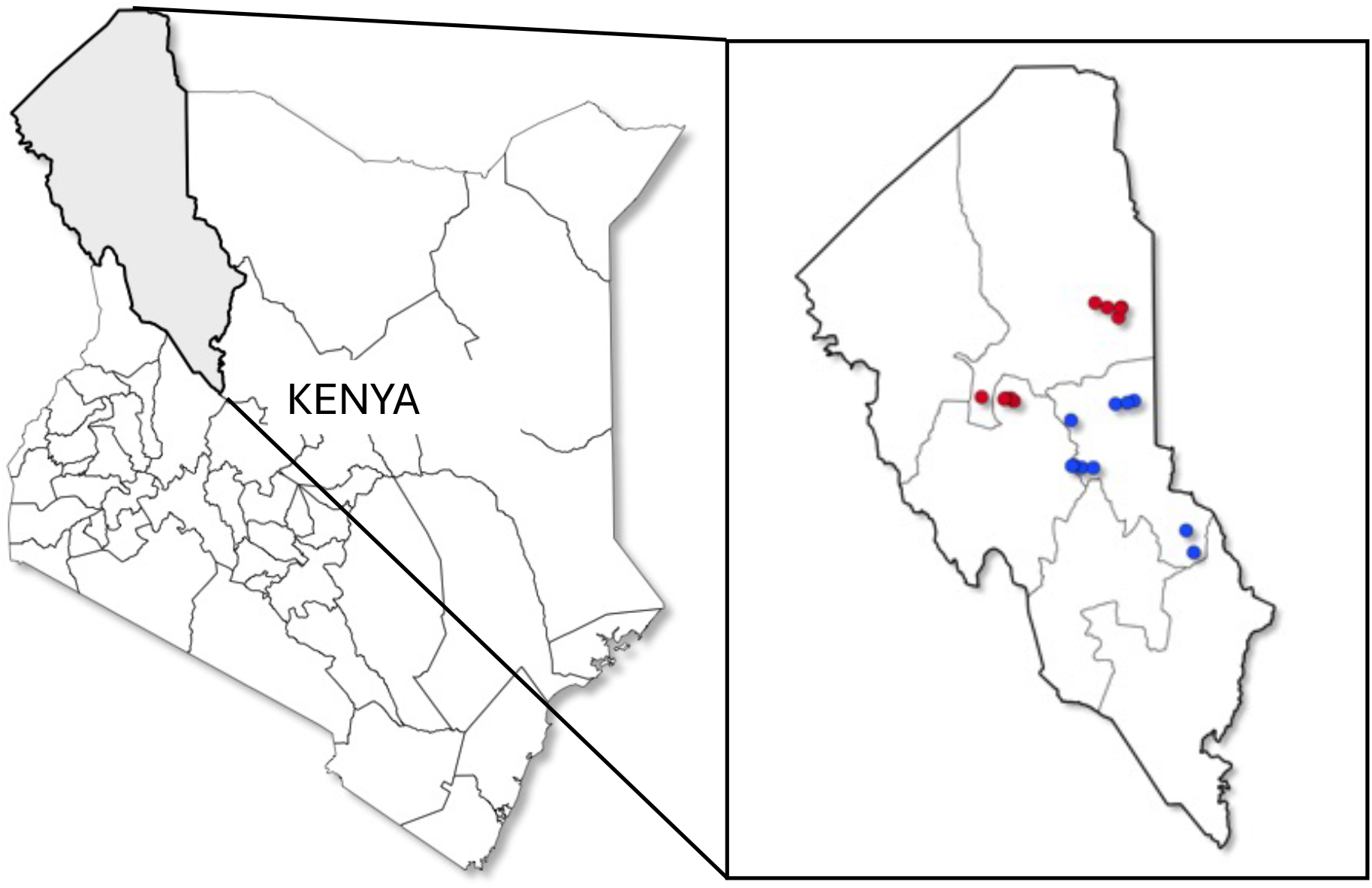
Map of the study villages in Turkana County, Kenya. Inset showing villages in Turkana Central subcounty (intervention area, blue dots) and Turkana North subcounty (comparison area, red dots).

**Figure 2:**
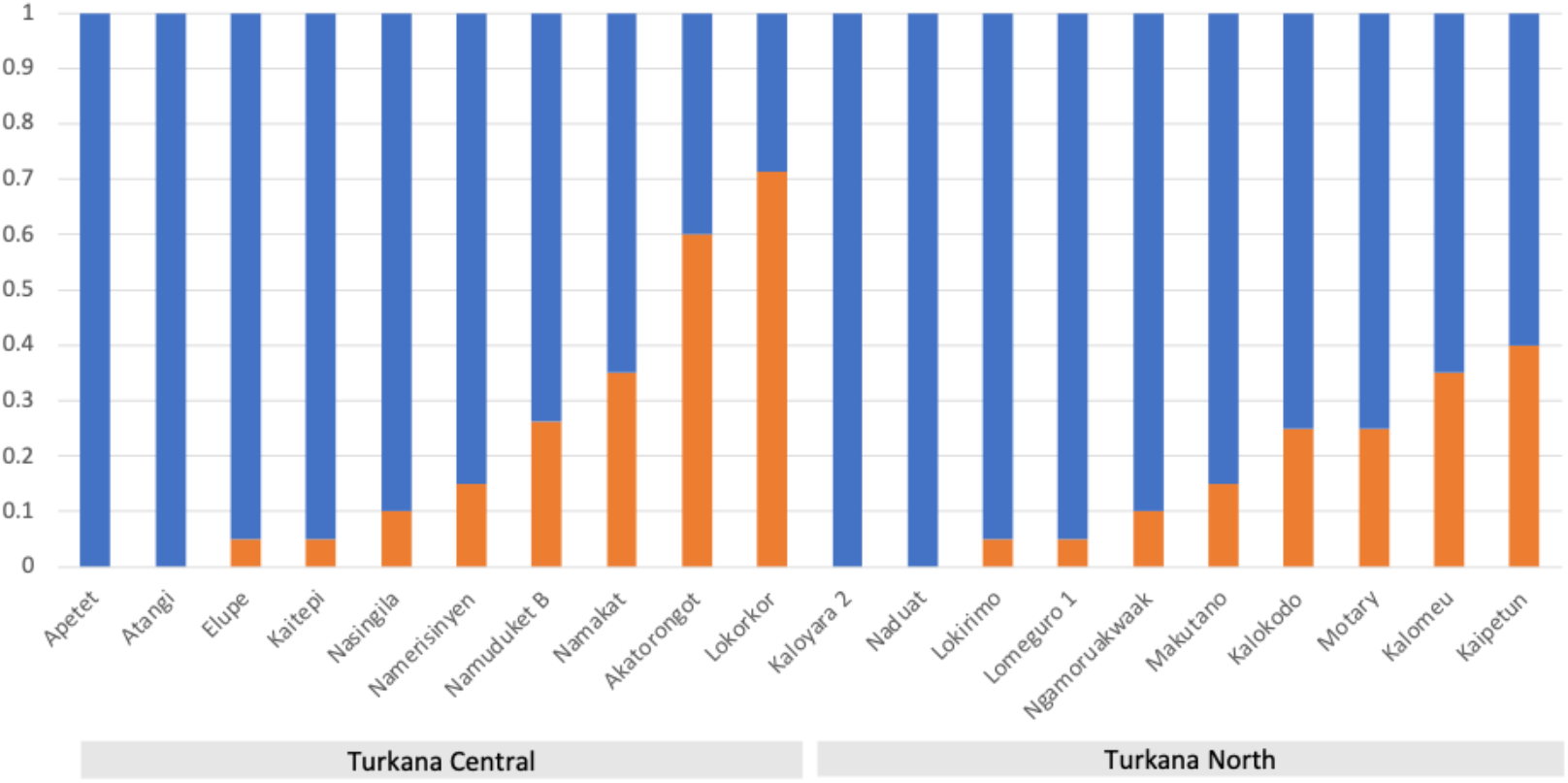
Parasite prevalence by village, measured by PCR, in all cohort children at enrollment

### SMC coverage in the Turkana Central cohort

All 198 children in Turkana Central were eligible for SMC. 143 of them (73%) received a full five cycles of SMC. Of those that did not receive all five cycles, 2 (1%) received no SMC at all and 39 (24%) received between 1-4 cycles. 14 children in Turkana Central were not found at endline and SMC coverage could not be determined exactly, but at least 10 of them had a minimum of one cycle of SMC according to records from weekly visits.

### Malaria episodes and protective efficacy of SMC

Malaria episodes in cohort children were recorded from the last day of the first SMC cycle until 28 days after the last cycle of SMC (June 18, 2024 – Nov 7, 2024). Twenty-two children (8 in Turkana North and 14 in Turkana Central) were lost to follow-up before the end of the study period and unable to be contacted at study close. Twenty-seven children traveled during the observation period and missed at least two weekly visits, but twenty-two of those were contacted at the final visit. In total, cohort children were followed for a 7585 child-weeks of observation (3779 in Turkana Central and 3806 in Turkana North). In the time period between the first SMC cycle and 30 days after the last cycle, 566 febrile events were documented, of which 207 were malaria episodes, giving a crude incidence of 87.5 malaria episodes per 100 person years at risk (PYAR) in Turkana Central and 202.7 per 100 PYAR in Turkana North. 159 children in Turkana Central remained malaria free throughout the study period compared to only 99 of their peers in Turkana North. After adjusting for child age, bednet use and cluster-level baseline malaria prevalence, the overall reduction in malaria cases attributed to SMC was 71% (Adjusted IRR 0.29, 95%CI:0.13-0.67, Table S2). An estimated 59% reduction in all fevers in the SMC group indicates that a high proportion of fevers are attributable to malaria (0.47, 95% CI: 0.31-0.72, Table S3). Figure 3 shows the number of cases per week in Turkana North versus Turkana Central over the study period.

**Figure 3.**
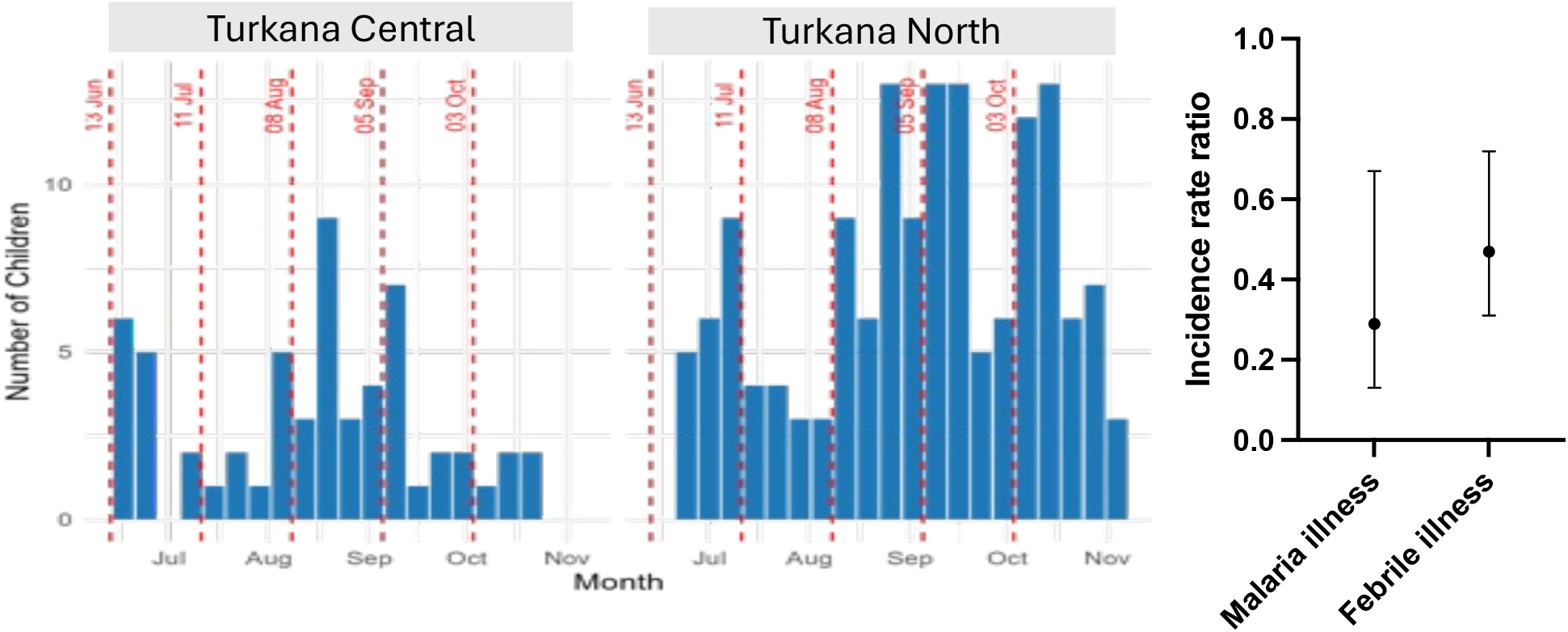
Malaria case count by study week in Turkana Central (left panel) and Turkana North (right panel). The dashed red lines indicate SMC distribution dates.

Children in the Turkana Central cohort who had incomplete SMC (fewer than 5 cycles) experienced more malaria episodes than children with complete SMC; 41 of 56 malaria episodes (73%) in SMC eligible children occurred in those who had incomplete SMC. Of the 11 children in Turkana Central who experienced more than one malaria episode in the study period, all but one had incomplete SMC (fewer than 5 cycles).

### Parasite resistance markers

351 participant DBS were collected by CHPs during the study period, 156 of which had parasites detected by PCR, 55/138 in Turkana Central and 101/213 in Turkana North. At least one of the three positions of interest amplified in 90 infections (*Pfdhfr* 59, *Pfdhps* 437 and 540, Table 3) and all three positions amplified in 68 infections. At least one mutation associated with resistance to SP was present in 54% of infections that amplified for all three markers tested (n=37/68). The majority of infections contained at least one wild-type allele out of the three positions tested. More infections had mixed alleles (both mutant and wildtype present in the infection) at each locus in Turkana Central than in Turkana North, indicating more multi-genomic infections. Although the prevalence of *Pfdhps* 437 mutation is significantly higher in Turkana Central, the distribution of the combinations of mutations was not significantly different between the groups (Table 4). The odds of harboring a mutant *Pfdhps* 437 or *Pfdhps* 540 allele increases with each additional SMC cycle received (Figure 4).

**Table 3:** Prevalence of wildtype, mutant type and mixed alleles in Turkana North and Turkana Central.

|  | Subcounty |  |  |  |
| --- | --- | --- | --- | --- |
|  | Central | North | Total | p-value* |
| *Pfdhfr* 59 (n=86) |  |  |  |  |
| Mixed | 5 (18.5%) | 6 (10.2%) | 11 (12.8%) | 0.500 |
| Mutant | 7 (25.9%) | 20 (33.9%) | 27 (31.4%) |  |
| Wild | 15 (55.6%) | 33 (55.9%) | 48 (55.8%) |  |
| *Pfdhps* 540 (n=70) |  |  |  |  |
| Mixed | 4 (17.4%) | 2 (4.3%) | 6 (8.6%) | 0.113 |
| Mutant | 5 (21.7%) | 7 (14.9%) | 12 (17.1%) |  |
| Wild | 14 (60.9%) | 38 (80.9%) | 52 (74.3%) |  |
| *Pfdhps* 437 (n=87) |  |  |  |  |
| Mixed | 7 (26.9%) | 12 (19.7%) | 19 (21.8%) | 0.030 |
| Mutant | 7 (26.9%) | 5 (8.2%) | 12 (13.8%) |  |
| Wild | 12 (46.2%) | 44 (72.1%) | 56 (64.4%) |  |

\*Chi square test

**Table 4:** Prevalence of combinations of mutant alleles and expected phenotype by subcounty.

|  | Subcounty |  |  |  |
| --- | --- | --- | --- | --- |
|  | Central | North | Total | p-value |
| N | 24 (32.4%) | 50 (67.6%) | 74 (100.0%) |  |
| Mutations<sup>1</sup> |  |  |  |  |
| *Pfdhps* A437G | 2 (8.3%) | 0 (0.0%) | 2 (2.7%) | 0.063 |
| *Pfdhps* A437G K540E | 3 (12.5%) | 3 (6.0%) | 6 (8.1%) |  |
| *Pfdhps* A437G, *Pfdhfr* C59R | 1 (4.2%) | 2 (4.0%) | 3 (4.1%) |  |
| *Pfdhps* A437G K540E, *Pfdhfr* C59R | 6 (25.0%) | 5 (10.0%) | 11 (14.9%) |  |
| *Pfdhfr* C59R | 1 (4.0%) | 13 (22.4%) | 14 (16.9%) |  |
| Wild-type | 10 (40.0%) | 27 (46.6%) | 37 (44.6%) |  |
| Resistance level<sup>2</sup> |  |  |  |  |
| 0 | 10 (41.7%) | 27 (54.0%) | 37 (50.0%) | 0.119 |
| 1 | 5 (20.8%) | 15 (30.0%) | 20 (27.0%) |  |
| 2 | 9 (37.5%) | 8 (16.0%) | 17 (23.0%) |  |

1. Based on previous studies in Kenya which found 100% prevalence of *Pfdhfr* N108S, N51I, and our pilot results on 10 samples which confirmed this, we assume that all parasites have these two mutations. 'Wild-type' indicates wild type at *Pfdhfr* amino acid 59 and 540 as well as *Pfdhps* 437. Six infections amplified as wild-type at *Pfdhfr* 59 and *Pfdhps* 437 but did not amplify at *Pfdhfr* 540 – were assumed to be wildtype.
2. Parasites that are wild-type at *Pfdhfr* amino acid 59 and 540 as well as *Pfdhps* 437 are here labeled 'wild type' with a resistance level of '0'. Parasites with mutations only at *Pfdhps* 437, *dhfr* 59 or *Pfdhps* 437 & *Pfdhfr* 59 are considered to have intermediate resistance (level 1). Parasites with *Pfdhps* K540E with or without any other mutations are considered to have higher levels of resistance (level 2).

**Figure 4:**
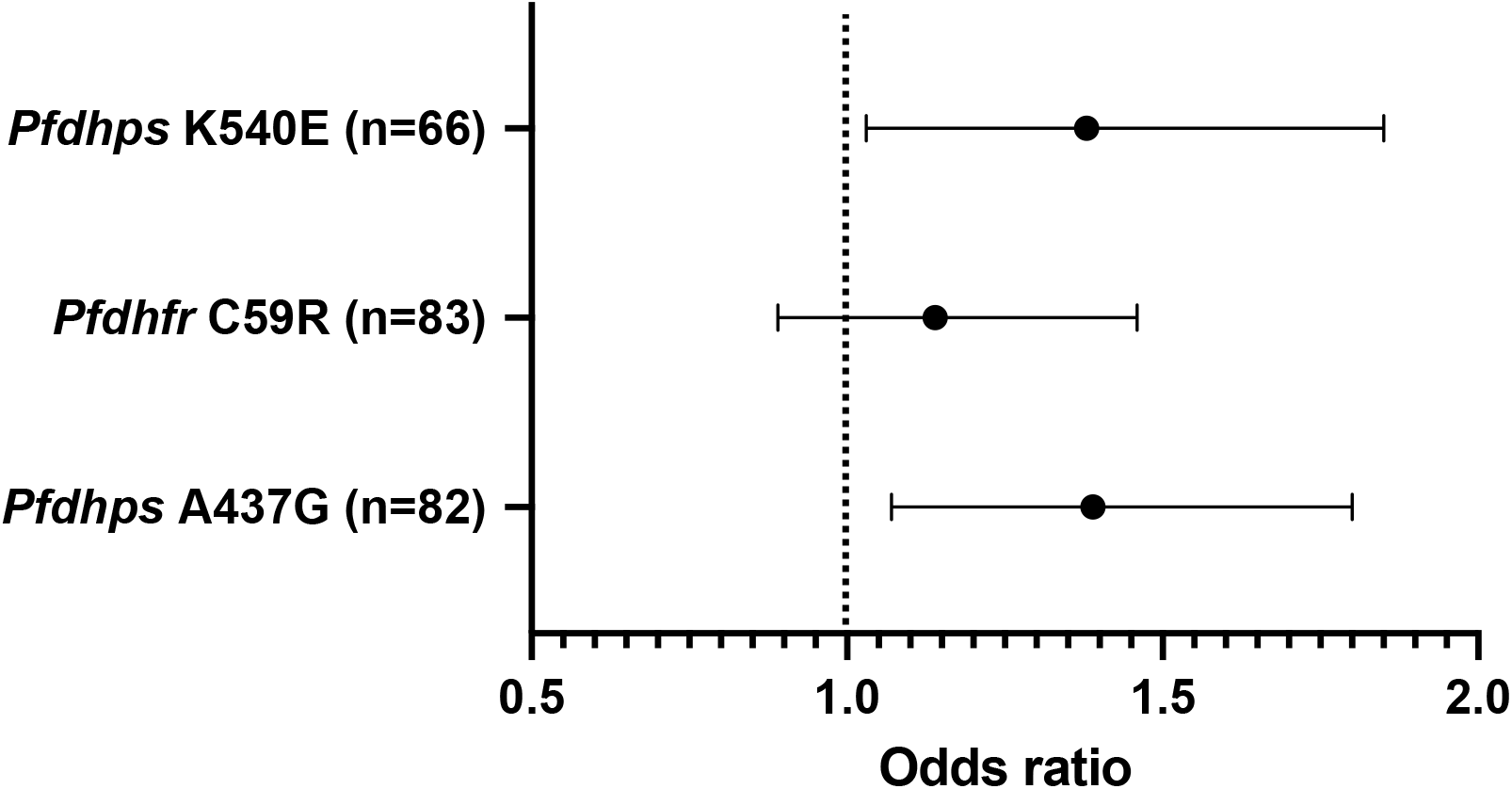
Unadjusted odds and 95% Confidence intervals of having an infection with a mutant allele at the indicated position for each additional cycle of SMC received between zero and five. Odds ratios are not adjusted for any covariates.

## Discussion

SMC is a highly cost-effective prevention strategy designed to reduce morbidity and mortality from malaria among children. However, experience with SMC in east and southern Africa is limited. In this study, SMC was successfully deployed within a unique community of pastoralists in northern Kenya and offered more than 70% protection against malaria illnesses during the five months of the program. The protection observed in the Turkana SMC program is lower than reported from the neighboring Karamoja study in Uganda, but generally agrees with randomized studies in West Africa (2) and estimates from routine implementation (6,36,37). Nearly three quarters of malaria episodes in the SMC cohort were among children who had incomplete SMC (<5 cycles), suggesting that the somewhat lower protection reported here is attributable to imperfect SMC adherence. In our cohort, despite the fact that children were in regular contact with the CHP, full adherence with all five cycles of SMC could only be confirmed in 78%.

Turkana Central is sparsely populated and villages are widely dispersed, with many being completely inaccessible during seasonal rains. Families are mobile, moving with livestock, but also for farming in floodplains and fishing. The remoteness and mobility of the population was expected to present a challenge for SMC coverage and indeed 12% of children missed contacts with the study team due to household movement. This is higher than we have been able to document in cross-sectional work and contributed to the suboptimal SMC uptake. We also observed that children who missed a cycle of SMC were very often sick at the subsequent cycle and again missed SMC. Although they would have received standard first-line treatment instead of SMC, artemether lumefantrine does not offer extended protection from reinfection, putting these children at risk of becoming infected again before the next cycle. This rotation of infection and ineligibility highlights the importance of timing the first cycle of SMC before significant transmission begins.

Parasite populations in East Africa have substantially higher levels and prevalence of antifolate resistance than those of West Africa (8), raising concerns about the efficacy of SPAQ for SMC in East Africa. Although SP has remained effective for protecting pregnant mothers, it is possible that pre-existing immunity may play a role in boosting the effect of the drug(38). The question of whether SPAQ would be less effective for preventing malaria in young, non-immune children exposed to drug resistant parasites has now been answered. Several studies, including ours, confirms high efficacy of SPAQ despite the predominance of drug-resistant parasites (14,39–41). To better understand the relationship between SMC and parasite resistance, previous studies have compared prevalence of resistance alleles before and after SMC in order to determine whether SMC may have increased the population prevalence of specific alleles(6,42,43). In our study, we focused on antifolate resistance alleles in clinical breakthrough infections during SMC to understand the relationship between SP resistance and effectiveness in preventing malaria. We detected a significantly higher occurrence of two *dhps* mutations in incident infections among children receiving SMC compared to incident infections in children not receiving SMC. Furthermore, children with more cycles of SMC had higher odds of harboring these mutations. One other study in Burkina Faso demonstrated selection for two *dhfr* alleles (*dhfr* 59R and 108N) in prevalent infections *during* SMC (44). Although it is not yet clear whether this small number of breakthrough infections may contribute to the spread of SP resistance, the observation that resistant parasites are enriched in, and may be increasing the risk of, breakthrough infections is important.

The relationship between the scale up of SMC with SPAQ and the change in prevalence of SP resistant parasites has also been explored in several studies from countries where SMC is routinely implemented at scale in West Africa. These studies typically measure changes in antifolate resistance alleles following SMC using repeated cross-sectional surveys of parasitemic but asymptomatic children or adults (6,42,43). Studies comparing infections across 2-5 years have not shown changes in antifolate resistance in the parasite population in SMC implementation areas (6,42), but studies that follow trends over longer periods in Africa indicates that antifolate resistance, especially pyrimethamine resistance, continued to expand over time following initiation of SMC (43). Although the relationship with SMC scale up cannot be confirmed, SMC is the only substantial source of selective pressure from SP. In East Africa, one study reported no difference in the prevalence of antifolate resistance mutations in children within and outside of SMC implementation areas in Uganda, although the baseline prevalence of resistance was very high which left little range to observe an increase (14). A major weakness of these ecological studies is the absence of information about the use of SMC at the individual level or enrollment of individuals not eligible for SMC.

Our study has limitations that should be considered when weighing the evidence. This was not a randomized study. Instead, we selected the comparison areas from which to draw the non-intervention villages to maximize similarity to the intervention area at baseline, reduce confounding, and therefore improve inference about the intervention effect. It is possible that weekly morbidity monitoring reduced eligibility for SMC as active monitoring is more sensitive than passive case detection. Children who have malaria illness on the day of SMC distribution or have taken antimalarials in the preceding two weeks are not eligible to receive SMC. The number of infections that we were able to type at each locus was small and therefore our comparisons are imprecise, although significant. Finally, we did not evaluate amodiaquine resistance in our cohort infections, although it is expected to be low in the region (45). Future studies should include genotyping of breakthrough infections at loci expected to confer resistance to SMC drugs.

Despite the high prevalence of parasites with mutations in *dhfr* and *dhps* linked to SP resistance, overall protection against malaria illness offered by SMC with SPAQ was high. This further highlights the value of expanding SMC to other regions experiencing highly seasonal transmission. However, given the scarcity in choice of chemoprevention drugs and the evidence of quintuple mutant infections in breakthrough infections of children receiving SMC, there is an urgent need for continuous monitoring of key mutations that may undermine SMC with SPAQ. This information is critical for countries considering adoption of SMC as an additional malaria intervention. Local levels of resistance remain important in informing parasite susceptibility and overall efficacy.

## Acknowledgements

We are deeply grateful for the families and CHPs who committed themselves to this work. We also acknowledge the enrolment team: Patrick Ejore, Zubeda Ewoi, Chuchu Gregory, Joseph Lobee, Lilian Lokitoe, Emmanuel Lopeto, Kaman Lowoi, Purity Musai, Joyce Nachipon, and Jeremiah Nokorot. We appreciate the contributions of Rebecca Lokwang, Lucy Abel, Shonde Akinola, Conde Mohamed Saran, Caroline Ngina, Tabitha Chepkwony, George Ambani and Emily Robie. Without the partnership and leadership of the Turkana County Health Team, this work would not have been possible.

## Data availability statement

Informed consent specifically included public data sharing and therefore data will be made available in a public repository after acceptance of this manuscript.

## Author statement

The authors declare they have no competing interests.

